# Effectiveness of a cash for school program on education and marriage among adolescent girls: A longitudinal intervention study in Lebanon

**DOI:** 10.1101/2025.08.26.25334430

**Authors:** Shatha Elnakib, Maia Sieverding, Zeinab Kammouni, Heba Dabliz, Karine Taha

## Abstract

**Introduction:** This study evaluates the effects of a three-year conditional cash transfer program on educational outcomes and child marriage among Lebanese, Syrian and Palestinian adolescent girls in Lebanon.

**Methods:** In the third year of the program, survey data were collected from households receiving a cash transfer conditional on adolescent girls’ school attendance and a comparison group of households. We conducted descriptive analysis and logistic regression models comparing outcomes, at endline, between girls in the intervention and comparison groups. Survey data on marriage and school dropout were also collected from all households of girls who were ever enrolled in the intervention or comparison groups over the three years of the program. Survival analysis was used to compare the transition to marriage and school dropout among girls who received cash transfers for different durations.

**Results:** Among year three participants, receiving cash transfers was associated with improved educational and attitudinal outcomes, but not reductions in early marriage. Comparing all program participants, receipt of two or more years of cash transfers was associated with significant reductions in the risk of early marriage but receipt of cash for one year only was not. Each additional year of cash receipt was associated with reductions in the risk of school dropout.

**Conclusion:** Our findings point to the promise of cash for education as an approach to improving school retention and reducing early marriage among adolescent girls in humanitarian settings, provided that cash transfers can be implemented for a sufficient duration of time.

**Implications and contributions:** This is one of the first studies to evaluate the impacts of a CCT program on early marriage in a humanitarian setting. Leveraging longitudinal data, our findings provide evidence on the promise of cash conditional on schooling to improve school retention and reduce early marriage among conflict-affected adolescent girls.

## Background

Despite its adverse impacts, child marriage remains prevalent in many parts of the world (1). UNICEF estimates that 650 million women globally were married as children, including 40 million in the Middle East and North Africa (MENA) (2). While much progress has been achieved in reducing the practice, the pace of decline remains too slow to meet the Sustainable Development Goal of ending child marriage by 2030 (2).

Child marriage is deeply rooted in sociocultural norms and propelled by poverty, lack of opportunities for girls, and low levels of agency (3). Amidst rising levels of armed conflict, there is debate over the impact of conflict and forced displacement on child marriage. Empirical evidence is mixed, suggesting that impacts on child marriage are context- and conflict-specific (4,5). Still, in the MENA region, high rates of child marriage have been documented among displaced and conflict-affected populations (4,6).

Lebanon currently hosts the highest number of refugees per capita globally, over half of whom are under 18 (7). These include 1.5 million Syrian refugees, who face risk factors for early marriage, including near-universal poverty and poor access to schooling, and around 200,000 Palestinian refugees who have experienced protracted displacement (8). Both refugee populations, as well as the Lebanese host population, have also been affected by a devastating economic crisis since 2019. Since the crisis began, school closures due to instability, COVID-19, and teacher strikes have further interrupted schooling. Pre-crisis estimates from 2015-2016 show 6% of Lebanese women aged 20– 24 married before age 18, compared to 12% of Palestinian refugees from Lebanon, 25% of Palestinian refugees from Syria, and 41% of refugees from Syria (9). In the context of heightened vulnerability post-crisis, identifying effective interventions to combat early marriage is all the more critical.

The role of school retention in preventing child marriage is well demonstrated (10–13). A recent review of interventions concluded that enhancing girls’ human capital and expanding their opportunities is the most effective approach to delaying marriage, with interventions that support girls’ education, particularly through cash or in-kind transfers, showing the most consistent success in preventing child marriage (10). Underpinning the success of conditional cash transfers (CCT) is their role in reducing the financial burden of education, making schooling a viable alternative to early marriage. By keeping girls in school, these programs increase girls’ future earning potential, raise the opportunity cost of child marriage, and empower girls with knowledge and confidence (10). These changes in turn slowly shape social norms that prioritize education over child marriage.

Despite a burgeoning evidence base, there is limited research on the impact of CCTs on child marriage in MENA, where entrenched gender norms and displacement may further entrench the practice. While cash programs have emerged as a preferred and popular intervention modality in humanitarian settings (14,15), their role in schooling retention and delaying marriage in such contexts remains under-explored. This paper evaluates a three-year conditional cash transfer program for girls’ education implemented by Anera, a Non-Governmental Organization (NGO), in North Lebanon. Using a quasi-experimental design, it assesses the program’s impacts on educational outcomes and marriage outcomes among Syrian, Palestinian and Lebanese adolescent girls.

## Methods

### Setting

Akkar, a governorate in northern Lebanon, was selected due to its high vulnerability and dense concentration of Syrian and Palestinian refugees and vulnerable Lebanese citizens. The region faces challenges of displacement, poverty, and underdevelopment, with strained infrastructure and services and growing social tensions due to the influx of refugees.

### Intervention

The Sawa Program was a three-year conditional cash transfer program seeking to address the financial barriers that hamper girls’ schooling and by extension reduce the risk of child marriage. The program ran for three academic years (AY), from fall 2021 through summer 2024. The primary intervention component was a conditional cash transfer for girls’ schooling, which was complemented by awareness activities at the community and household levels.

In Year 1 of the program (AY 2021–22), Syrian and Lebanese girls received two payments: $150 as the first installment and $100 as the second, with an additional $50 provided to those enrolled in Grades 9 and 12 to support exam-related needs. These payments were distributed mid-year and toward the end of the academic year, aligning with key educational milestones to encourage continued attendance and exam preparation.

Palestinian girls received a one-time payment of $200, disbursed in the second half of the school year. This approach was informed by consultations with caregivers and school staff, which highlighted a preference for a single, consolidated transfer during the initial year of program implementation in schools serving Palestinian communities.

In Year 2 (AY2022–23), all eligible girls received increased support, with a first payment of $250, followed by a second payment of $200. In Year 3 (AY2023–24), due to funding constraints, the financial assistance was reduced to a total of $200, distributed in two separate installments. In all three years, and for girls of all nationalities, receipt of the cash transfers was conditional on regular school attendance and continued enrollment.

### Participants

The program was originally designed as a two-year cash transfer. Over AY 2021-22 and 2022-23, the program enrolled a total of 948 intervention participants (Figure 1). A comparison group of 279 participants (split over the two years) was established. The program was then renewed for a third year (AY 2023–24) and included 378 continuing participants from Years 1 and 2, along with 340 newly identified girls, totaling 718 intervention participants. An additional 357 girls were newly recruited to form the Year 3 comparison group. Quota sampling was used to enable a participant breakdown that reflects the burden of child marriage by nationality, resulting in approximately 10% of the sample being Lebanese, 20% Palestinian, and 70% Syrian. Supplementary Table S1 provides the distribution by intervention status, nationality, and year.

**Figure 1:**
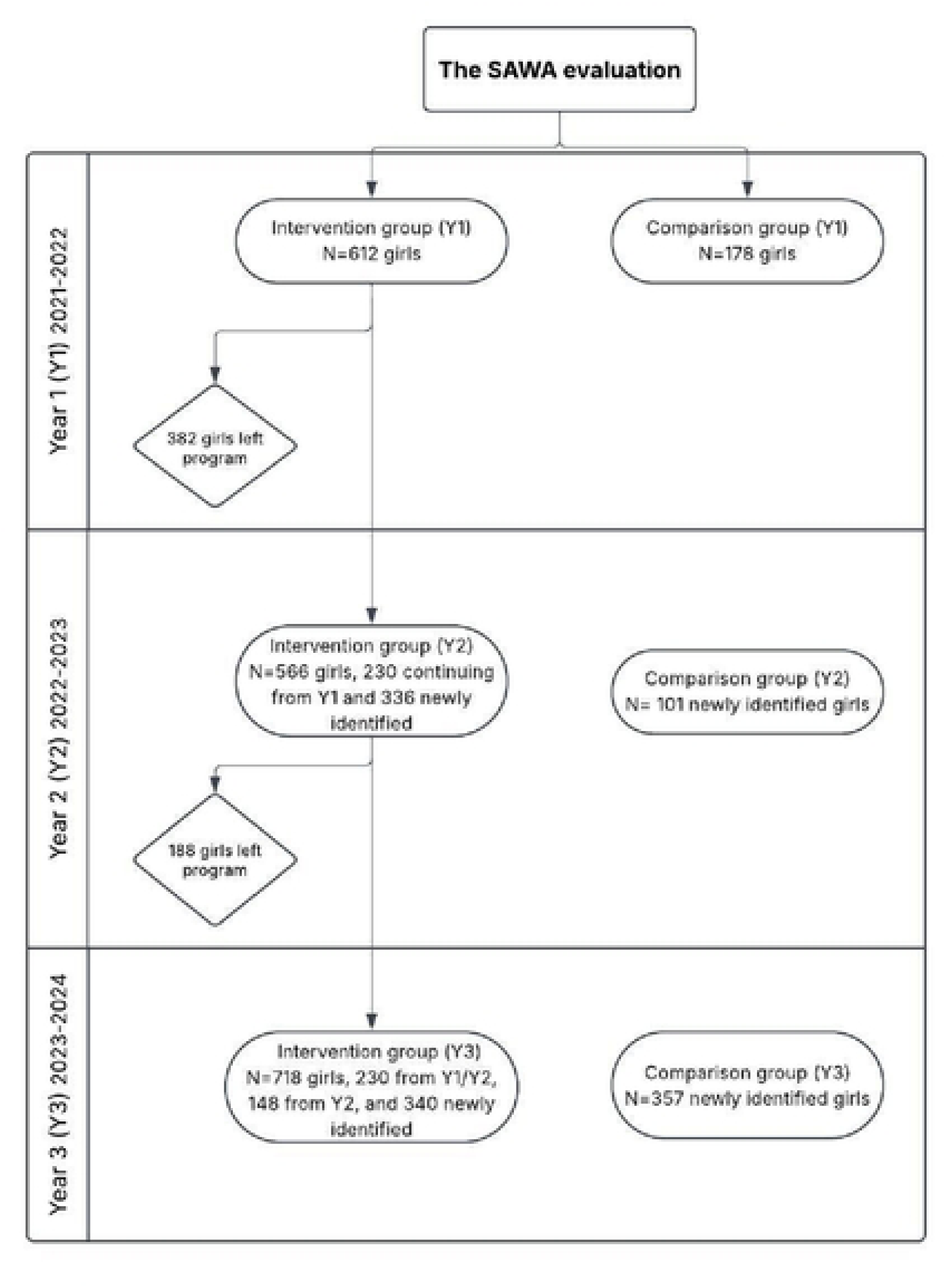
Participants in the SAWA program by year

In Year 1, participants were selected based on a vulnerability assessment measuring demographic, economic, and shelter-related risks. Of 2,000 households assessed, Anera selected 612 girls with the highest scores to receive the intervention and matched them with comparison participants with similar vulnerability scores. In Years 2 and 3, schools nominated vulnerable girls, from whom participants were randomly assigned to the intervention or comparison group.

Eligibility criteria included enrollment in public, private, or UNRWA schools in Years 1 and 2; private schools were excluded in Year 3. Girls were aged 13–16 in the first two years, with the upper age limit extended to 17 in Year 3 to accommodate continuing participants. While comparison participants did not receive cash for schooling, they were referred to other Anera programs offering in-kind support, such as food parcels and medical supplies.

### Data collection

Our analysis is based on data collected during Year 3 of the program. Baseline data for Year 3 participants were collected in person between 1 December 2023 and 30 January 2024, before their participation in the program and was delayed due to delays in the start of the school year. The baseline survey captured girls’ age, school status, and proxies for household economic conditions. Endline data were collected by phone from August to September 2024, following the academic year and final exams. Core outcomes were school retention, attendance, self-reported academic performance, and marital status, and official school grades obtained from schools were merged when available. Girls were also asked directly about their knowledge and attitudes toward early marriage. Complete baseline and endline data were obtained for 1,033 girls; 702 in the intervention group and 331 in the comparison group.

In November 2024, a brief phone survey was conducted with caregivers of girls from Years 1 and 2 who were no longer enrolled in Year 3, capturing current school enrollment and marital status, including dropout and dates of marriage when relevant. Of the 1,924 girls ever enrolled, data were obtained for 1,800 through either the Year 3 endline or the follow-up survey.

### Outcomes

Our primary outcome is a binary indicator for whether the girl had ever been married (yes = 1). Given the low incidence of marriage among Year 3 participants, we examined a second outcome for whether the girl had ever been married or engaged. For those ever married, age at marriage was calculated by subtracting date of birth from date of marriage. The key educational outcome is school retention. For Year 3 participants, this is defined as a binary indicator for whether the girl completed the full 2023–24 academic year and sat for final exams. For the full study sample, we calculated age at school dropout for girls no longer enrolled at endline. For both marriage and dropout, dates often clustered on the first of the month but this should not substantially affect results.

Additional outcomes, analyzed only for Year 3 participants, include school attendance (self-reported as >90%, 74–90%, or <75%), and academic performance based on school-reported final grades. We construct a continuous grade (out of 100) and a binary indicator for passing the school year (grade >50%, per Lebanese standards). The latter definition includes some students for whom only pass/fail status was provided. We also assessed families’ intent to re-enroll the girl for the 2024–25 school year, acknowledging that the survey preceded the disruptions caused by the 2024 conflict.

For families of unmarried girls, we asked whether the girl had received a marriage proposal and, if yes, constructed a binary indicator for whether the proposal was rejected (=1) or still under consideration. We also examined whether girls could identify at least one negative consequence of early marriage (health, social, or economic), and analyzed attitudes using agreement with the statements “Puberty means a girl is ready for marriage” and “Marriage is protection for a girl.” Responses were recorded as binary indicators, distinguishing between disagreement versus agreement or neutrality.

### Statistical analysis

We compared baseline characteristics of Year 3 intervention and comparison groups using chi-squared tests for categorical variables and t-tests for continuous variables. Characteristics included nationality, age, school type (public vs. UNRWA for Palestinians), disability/chronic illness, employment, dwelling type (apartment, informal tented settlement, or other), and a household vulnerability index. The index was constructed via factor analysis of five baseline variables: household income, cash assistance receipt (other than the program), dwelling type, home ownership, and household debt.

Endline outcomes were compared between groups using the same statistical tests. While this unadjusted analysis does not control for baseline differences, it allows us to examine behavioral and attitudinal outcomes such as school re-enrollment intentions and responses to marriage proposals, not measured at baseline.

For key outcomes, we estimated logistic regression models with intervention status as the main predictor. Models controlled for girls’ age and household vulnerability; models for marriage also included nationality, while school outcome models included school type. Although this approach does not fully eliminate baseline imbalances, it is an improvement over simple group comparisons. Behavioral, knowledge, and attitudinal outcomes were excluded from regression analysis due to the absence of baseline data. All regressions clustered standard errors at the household level.

To analyze marriage and school dropout across all three program years, we used survival analysis techniques to account for censored observations. Kaplan-Meier failure curves were constructed by duration of cash transfer receipt. Cox proportional hazard models were then estimated using an ordinal variable for number of years of support, controlling for age and nationality. Due to small sample sizes, girls receiving two and three years of support were grouped, as were Lebanese and Palestinian participants in the marriage models. All analyses were conducted in Stata 16.

### Ethics approval

Oral consent was solicited from caregivers prior to any data collection activities and oral assent was sought from girls who were administered survey questions (those in Year 3). This study received ethics approval from the American University in Beirut Institutional Review Board (IRB# SBS-2024-0331).

## Results

### Characteristics of Year 3 intervention and comparison groups

Girls in the Year 3 intervention and comparison groups differed on most characteristics at baseline (Table 1). Girls in the intervention group were slightly older (p<0.001) and half as likely to be working as girls in the comparison group (p<0.001). Girls in the intervention group were more likely to live in apartments as opposed to an ITS or another form of sub-standard housing (p<0.001). They were also more likely to fall in the middle tertile of the vulnerability index, whereas girls in the comparison group were overrepresented among the most vulnerable (p=0.004).

**Table 1:** Characteristics of Year 3 intervention and comparison group participants.

|  | Comparison<br>N=331 | Intervention<br>N=702 | Total<br>N=1,033 | p-value* |
| --- | --- | --- | --- | --- |
| **Nationality** |  |  |  | 0.63 |
| Lebanese | 37 (11.2%) | 65 (9.3%) | 102 (9.9%) |  |
| Palestinian | 67 (20.2%) | 144 (20.5%) | 211 (20.4%) |  |
| Syrian | 227 (68.6%) | 493 (70.2%) | 720 (69.7%) |  |
| **Age** | 14.1 (1.3) | 14.8 (1.4) | 14.6 (1.4) | <0.001 |
| **School type** |  |  |  | 0.70 |
| Public | 263 (79.5%) | 565 (80.5%) | 828 (80.2%) |  |
| UNRWA | 68 (20.5%) | 137 (19.5%) | 205 (19.8%) |  |
| **Disability/chronic illness** | 38 (11.5%) | 48 (6.8%) | 86 (8.3%) | 0.012 |
| **Girl employed** | 65 (19.6%) | 62 (8.8%) | 127 (12.3%) | <0.001 |
| **Dwelling type** |  |  |  | <0.001 |
| Other | 72 (21.8%) | 123 (17.5%) | 195 (18.9%) |  |
| ITS | 93 (28.1%) | 113 (16.1%) | 206 (19.9%) |  |
| Apartment | 166 (50.2%) | 466 (66.4%) | 632 (61.2%) |  |
| **Household vulnerability score** |  |  |  | 0.004 |
| Most vulnerable | 127 (38.4%) | 223 (31.8%) | 350 (33.9%) |  |
| Middle tercile | 88 (26.6%) | 259 (36.9%) | 347 (33.6%) |  |
| Least vulnerable | 116 (35.0%) | 220 (31.3%) | 336 (32.5%) |  |
| **Previous years in intervention** |  |  |  | - |
| No | - | 331 (47.2%) |  |  |
| Yes | - | 371 (52.8%) |  |  |

Notes: ITS = informal tented settlement
\*p-values are based on a chi-squared test for categorical variables and a t-test for continuous variables

### Year 3 intervention outcomes

At endline, there was no significant difference in the prevalence of early marriage or engagement between girls in the Year 3 intervention and comparison groups (p=0.67), with such events rare overall (Table 2). One girl in the comparison group was engaged and one was married; in the intervention group, one girl was married and five were engaged. Results remained nonsignificant when analyzing marriage and engagement separately.

**Table 2:**
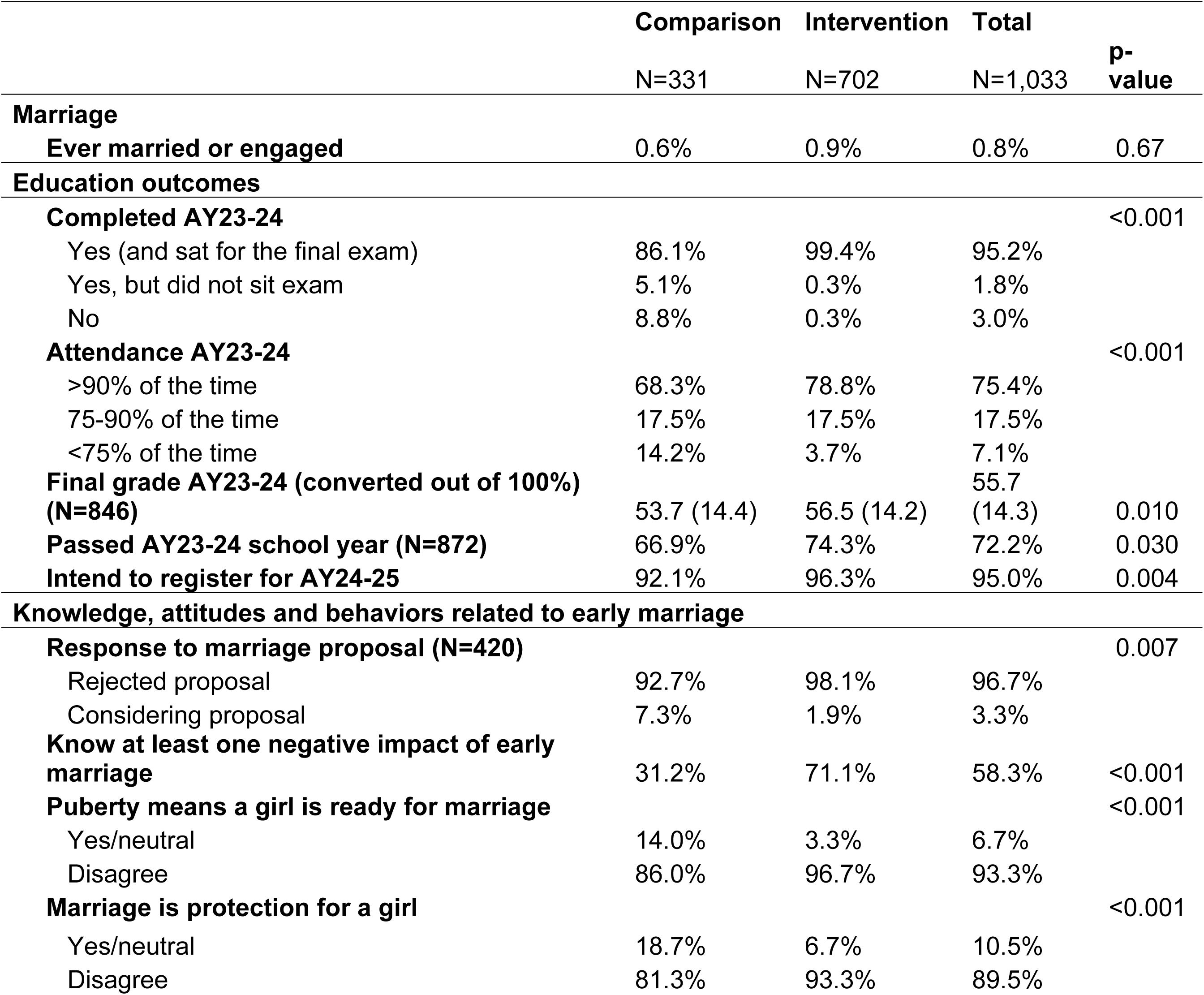
Outcomes among Year 3 intervention and comparison groups at endline.

By contrast, the intervention group outperformed the comparison group across all educational outcomes for the 2023–24 academic year. Intervention participants were significantly more likely to complete the school year (p<0.001) and attend regularly (p<0.001). They also had higher average final grades (p=0.010) and were more likely to pass the year (p=0.030). Additionally, caregivers of girls in the intervention group were more likely to report plans to re-enroll them for the 2024–25 school year (p=0.004).

Logistic regression models adjusting for age, nationality, school type, and vulnerability confirmed that intervention status was associated with improved educational outcomes but not with differences in marriage or engagement (Supplementary Table A2).

Significant differences were also observed in knowledge, attitudes, and behaviors related to early marriage (Table 2). Overall, 41% of girls had received a marriage proposal. Among these, the rate of rejecting the proposal was higher in the intervention group than the comparison group (p=0.007). Girls in the intervention group were also significantly more likely to identify at least one negative consequence of early marriage (p<0.001). While attitudes against child marriage were widespread (over 95% of girls in both groups reported that marriage under age 18 was unacceptable - data not shown), intervention participants were significantly more likely to disagree with the statements that reaching puberty means a girl is ready for marriage (p<0.001) and that marriage provides protection for girls (p<0.001).

### Long-term follow-up with all program participants

Table 3 presents the characteristics of all girls over Years 1-3. One-third never received the cash transfer, having participated only as part of the comparison group. Nearly half received the transfer for one year, with smaller proportions receiving it for two or three years. Participants were, on average, 14.0 years old at enrollment and 15.7 at the time of the survey. Among the 1,773 girls with complete survey data, 4.8% had ever been married and 6.8% had ever been married or engaged. Nearly one-quarter had left school by the time of the survey. These outcomes were closely linked: almost all girls who had ever been married (92.9%) and over half of those who were engaged (58.3%) had also dropped out of school (data not shown).

**Table 3:**
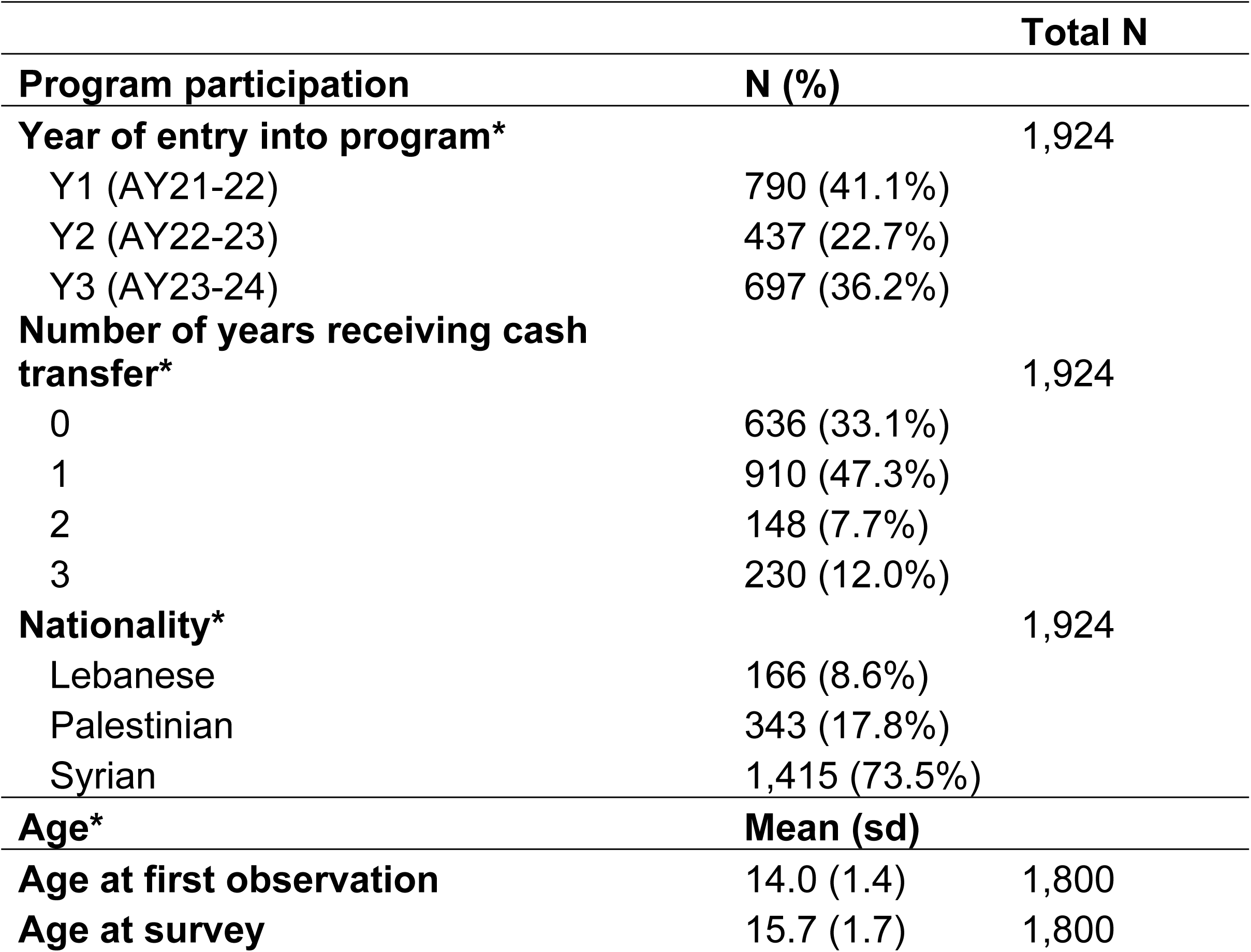
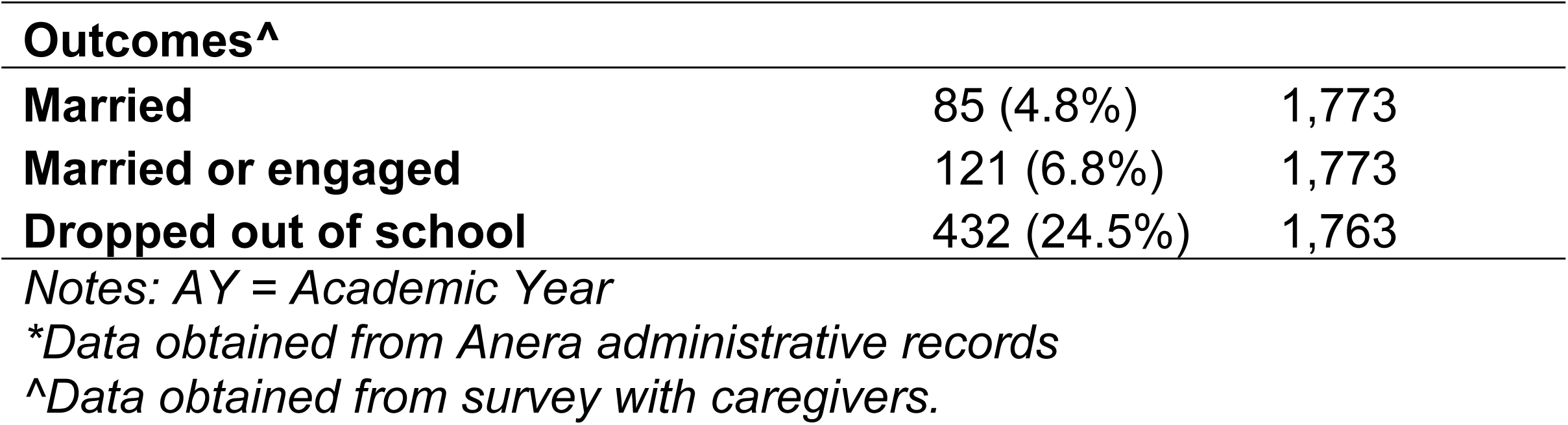
Characteristics of participants over the three years of the program, AY2021-22 through AY2023-24.

### Early marriage

Survival analysis suggests that the intervention significantly delayed marriage among girls who received the cash payments for two or three academic years. By contrast, girls who received the cash payments for only one academic year had a pattern of early marriage very similar to those who did not receive the cash payments at all (Figure 2, Panel A). Results using marriage or engagement as the outcome were substantively identical (Supplementary Figure F1). Results of the Cox proportional hazard models confirm that receiving 2-3 years of cash payments was associated with a significantly lower hazard of marriage, even when controlling for age and nationality, whereas receiving only one year of cash payments was not (Table 4, Col. 1).

**Figure 2:**
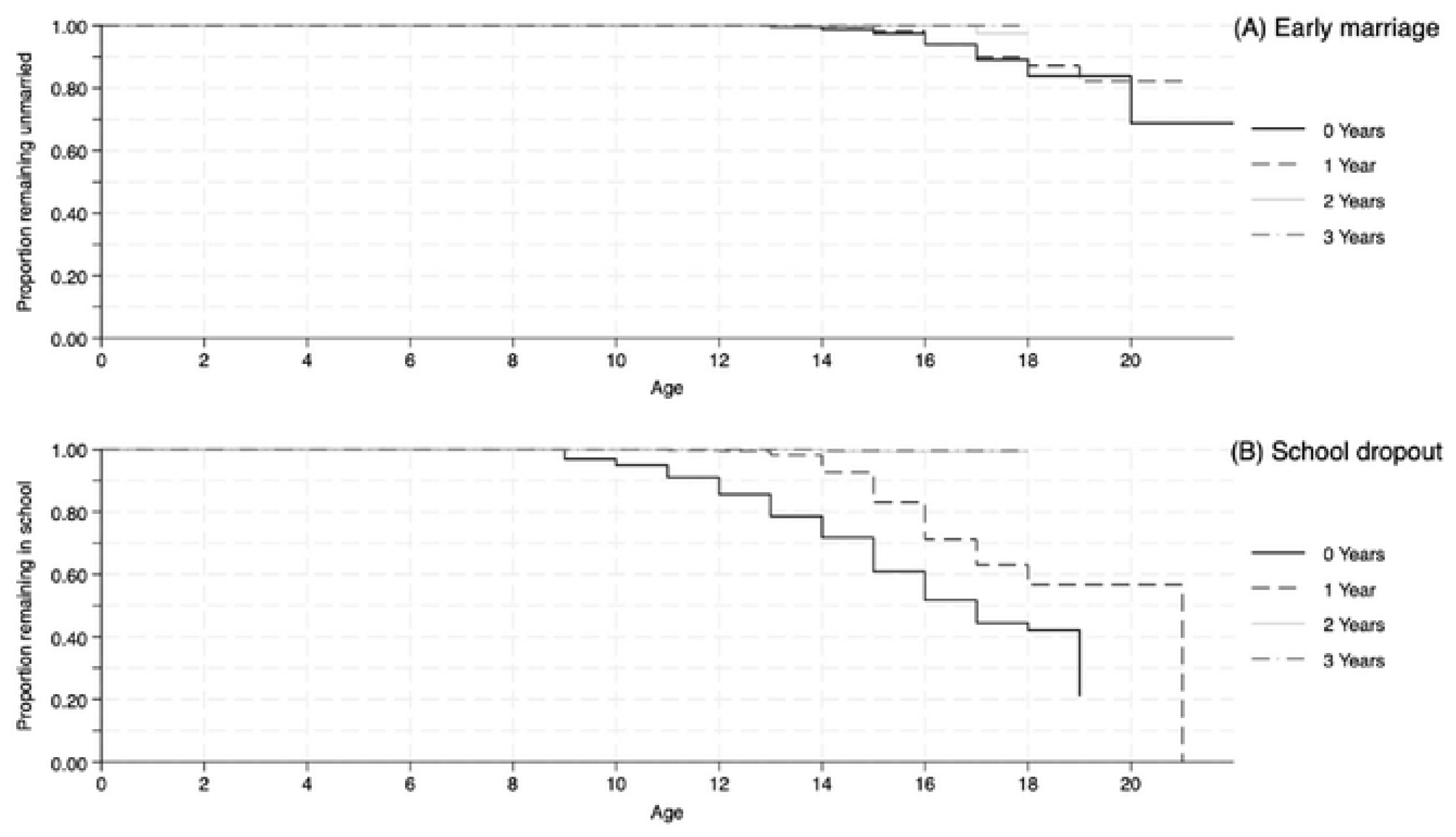
Survival distribution of school dropout and marriage

**Table 4:**
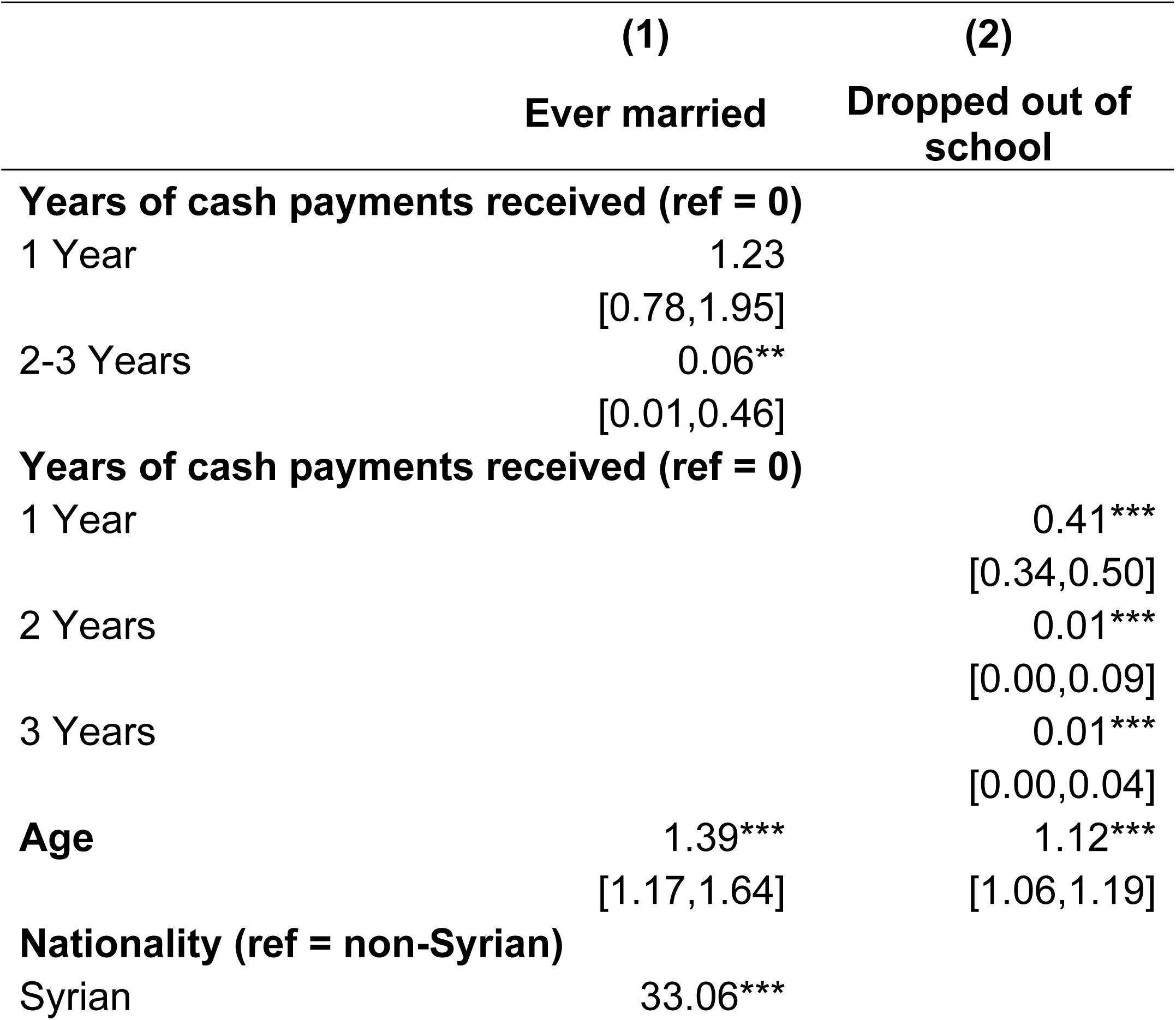
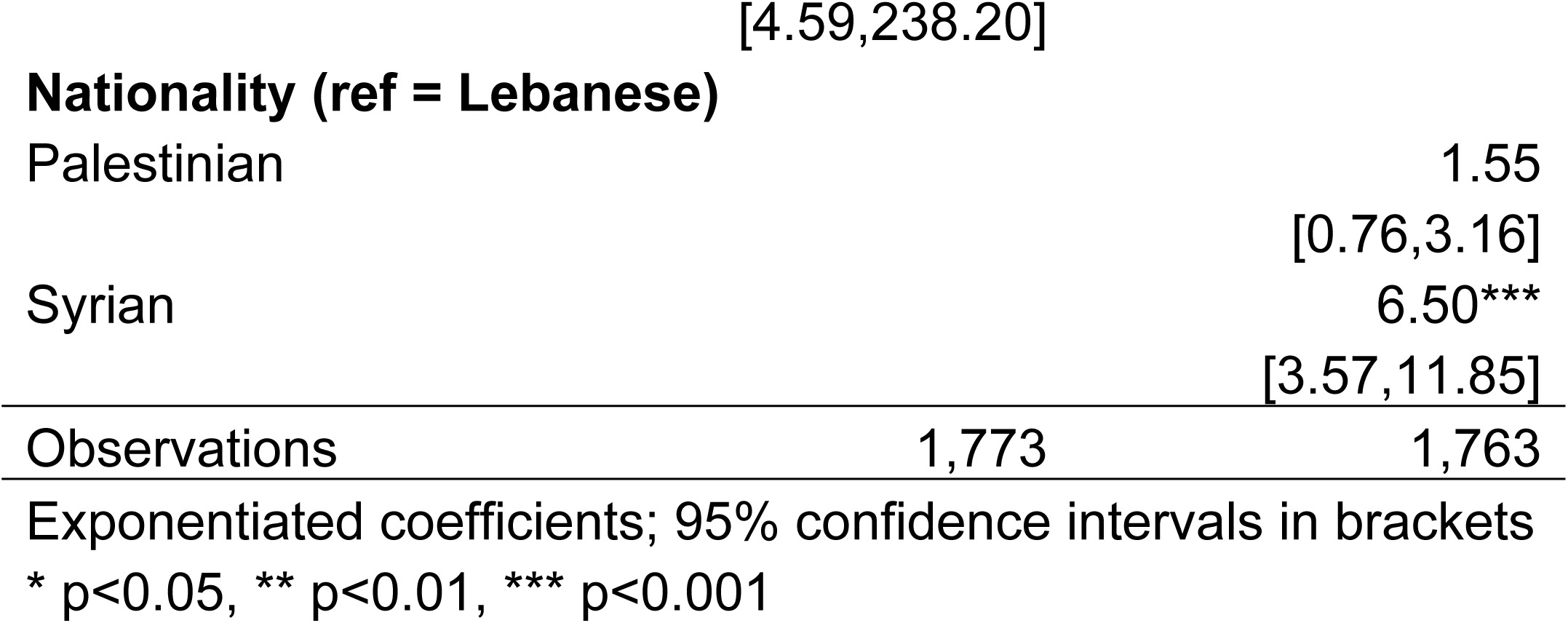
Cox proportional hazard models for early marriage and school dropout, Y1-Y3 participants (hazard ratios)

### School retention

As with early marriage, there was almost no dropout among girls who received the cash payments for two or three years (Figure 2, Panel B). However, there was also a significant difference between girls who did not receive cash payments at all and those who received payments for only one year. Among the former, the 25^th^ percentile for school retention was age 14 and among the latter, it was 16. The Cox proportional hazard model confirms this finding (Table 4, Col. 2). Receiving one year of cash was associated with significantly lower odds of school dropout. Receiving two or three years of cash transfers was significantly associated with a lower hazard of dropout relative to the comparison group and to receiving one year of cash transfer. However, the difference between receiving two vs. three years of transfers was not significant. As with early marriage, age and Syrian nationality were significantly associated with a higher hazard of dropout.

## Discussion

Our study provides an in-depth analysis of the short- and longer-term impact of conditional cash transfers on girls’ educational outcomes as well as on the more distal outcome of early marriage. In Year 3, girls who received cash were significantly more likely to stay in school, attend regularly, pass exams, and plan to re-enroll the following year. They were also more likely to reject marriage proposals and showed greater awareness of the risks of early marriage. However, one year of support – while effective for improving education outcomes – was not sufficient to significantly reduce rates of early marriage among girls already enrolled in school. Over a longer follow-up period, girls who received cash transfers for two or more years were significantly less likely to marry early and had lower school dropout rates compared to those who received one year of support or none at all. Each additional year of cash assistance helped reduce dropout compared to no support, but the benefits were substantially greater with sustained investment.

Our findings align with other evaluations of the impact of cash for education on schooling outcomes. Existing studies on CCTs have documented their effectiveness in increasing school enrollment, attendance, and sometimes progression, while impacts on learning outcomes have been more mixed. For example, a review by Baird et al. found that CCTs significantly increase school participation among children and adolescents (16), and a more recent meta-analysis found that CCTs can increase enrollment, with larger effects for secondary school students (17).

While we find important impacts on school retention and attendance, impacts on grades were less significant. Some studies of CCTs have documented modest or mixed improvements in grades, while others suggest little or no effect (16,18,19). This is not particularly surprising as learning tends to rely heavily on supply-related factors such as quality of teaching, school infrastructure and the learning environment. Our findings add evidence to the argument that the very act of staying in school may prevent marriage, as those two phenomena tend to be mutually exclusive. Indeed, our experience in Lebanon corroborates this assumption, as we were consistently told by school administrators that girls who get married are not allowed back into school, out of concerns that they would “corrupt” their unmarried peers.

Our finding that receipt of cash transfers for one year did not translate into a relative reduction in child marriage, but that a longer duration did, has important implications for the design of CCTs with conditionalities around school enrollment and the sustainability of impacts after cash transfers end. Some studies evaluating cash programs have found evidence of long-term effects. For example, Baird et al. find that conditional cash transfers had enduring impacts on certain aspects of human capital, such as education, which once acquired, did not disappear after the transfer ended (11). Another study in Kenya evaluating a two-year “cash plus” program for adolescent girls found sustained impacts on marriage and fertility outcomes six years after the intervention began, extending into late adolescence and early adulthood (20). These findings underscore the impacts of human capital investments on child marriage over the long term --but only if they meaningfully alleviate the economic pressures that drive early marriage. In our study, a one-year cash transfer was insufficient to disrupt these pressures, whereas support sustained over two or more years had a measurable effect. Indeed, a recent review undertaken by Girls Not Brides concluded that programs that are conducted with a regularity and predictability, and that extend over a longer period, are more likely to mitigate the economic pressures that contribute to child marriage (21). Ensuring sufficient duration of cash transfer programs is therefore crucial in addressing child marriage, particularly in humanitarian settings where implementation conditions and funding are sometimes precarious and patchy.

Our findings should be interpreted in light of several limitations. Over the course of the program, the country experienced several bouts of increased economic and political instability which led to repeated school closures and interruptions in program implementation, likely attenuating the intervention’s impact. Moreover, program components, including the amount of cash assistance, were not fully standardized over time or across subgroups due to funding constraints and variability in participants’ vulnerability profiles. Additionally, despite efforts to match comparison and intervention groups, baseline differences in Year 3 suggest the potential for selection bias in the observed effects.

These limitations notwithstanding, this is one of the first studies to evaluate the impacts of a CCT program on early marriage in a humanitarian setting. The fact that we were able to follow participants several years after entering the program also provides important evidence on longer-term impacts, particularly in a context marked by instability. Our findings point to the promise of cash for education as an approach to improving school retention and reducing early marriage among adolescent girls affected by conflict, provided that cash transfers can be implemented for a sufficient duration of time.

## Data Availability

Upon acceptance, the data will be uploaded onto Figshare

## Declarations

## Disclosure of potential conflicts, real and perceived, for all named authors

There are no real or perceived conflicts of interest in undertaking or publishing this research.

## List of abbreviations

AY: Academic years
CCT: Conditional cash transfers
MENA: Middle East and North Africa
NGO: Non-Governmental Organization

## Authors’ contributions

HD and SE supervised the study. SE and MS conducted data analysis and led manuscript writing. HD, KT, and ZK contributed to data analysis and write-up of study findings. ZK and HD supported data management and quality assurance. HD, SE, and KT conceptualized

## References

1. Malhotra A, Elnakib S. Evolution in the evidence base on child marriage. 2021;

2. UNICEF. UNICEF DATA. 2018 [cited 2021 Jan 1]. Child Marriage: Latest trends and future prospects. Available from: https://data.unicef.org/resources/child-marriage-latest-trends-and-future-prospects/

3. Psaki SR, Melnikas AJ, Haque E, Saul G, Misunas C, Patel SK, et al. What Are the Drivers of Child Marriage? A Conceptual Framework to Guide Policies and Programs. J Adolesc Health. 2021 Dec;69(6S):S13–22.

4. Shatha Elnakib et al. Investigating incidence, correlates, and consequences of child marriage among Syrian refugees residing in the south of Lebanon: a cross-sectional study. Journal of Adolescent Health.

5. Neal S, Stone N, Ingham R. The impact of armed conflict on adolescent transitions: a systematic review of quantitative research on age of sexual debut, first marriage and first birth in young women under the age of 20 years. BMC Public Health [Internet]. 2016 Mar 4 [cited 2019 Oct 6];16. Available from: https://www.ncbi.nlm.nih.gov/pmc/articles/PMC4779256/

6. UNICEF. Child marriage in the Middle East and North Africa. UNICEF, www.unicef.org/mena/media/1786/file/MENA-ChildMarriageInMENA-Report_pdf.pdf (accessed 11 October 2020). 2017;

7. UNHCR Lebanon. UNHCR Lebanon. [cited 2025 Mar 28]. UNHCR Lebanon. Available from: https://www.unhcr.org/lb/about-us/unhcr-lebanon-glance

8. Jad C, Hala G, Rima H, Sari H, Nadine S, Nisreen S, et al. Socio-Economic Survey of Palestine Refugees in Lebanon. Beyrouth, American University of Beirut, European Union, Unrwa. 2010;

9. UNICEF Middle East and North Africa Regional O_ce. Lebanon Country Brief UNICEF Regional Study on Child Marriage In the Middle East and North Africa [Internet]. [cited 2025 Mar 28]. Available from: https://www.unicef.org/mena/media/1806/file/MENA-CMReport-LebanonBrief.pdf%20.pdf

10. Malhotra A, Elnakib S. 20 Year of the Evidence Base on What Works to Prevent Child Marriage: A Systematic Review. Journal of Adolescent Health. 2020;

11. Baird S, McIntosh C, Özler B. When the money runs out: do cash transfers have sustained effects on human capital accumulation? The World Bank; 2016.

12. Lee-Rife S, Malhotra A, Warner A, Glinski AM. What Works to Prevent Child Marriage: A Review of the Evidence. Studies in Family Planning. 2012;43(4):287–303.

13. Wodon Q, Male C, Nayihouba A, Onagoruwa A, Savadogo A, Yedan A, et al. Economic impacts of child marriage : global synthesis report. MINISTERIO DE EDUCACIÓN [Internet]. 2017 Jun [cited 2020 Jan 20]; Available from: http://repositorio.minedu.gob.pe/handle/MINEDU/5588

14. International Rescue Committee (IRC). Why is cash assistance a critical form of humanitarian aid? [Internet]. [cited 2025 Mar 28]. Available from: https://www.rescue.org/article/why-cash-assistance-critical-form-humanitarian-aid

15. Barder O, Blattmann C, Cameron L, Egeland J, Elmi M, Faye M, et al. Doing cash differently: how cash transfers can transform humanitarian aid. ODI Rep (September). 2015;44.

16. Baird S, Ferreira, Francisco H.G., Özler, Berk, and Woolcock M. Conditional, unconditional and everything in between: a systematic review of the effects of cash transfer programmes on schooling outcomes. Journal of Development Effectiveness. 2014 Jan 2;6(1):1–43.

17. García S, Saavedra JE. Educational Impacts and Cost-Effectiveness of Conditional Cash Transfer Programs in Developing Countries: A Meta-Analysis. Review of Educational Research. 2017 Oct 1;87(5):921–65.

18. Glewwe P, Muralidharan K. Improving Education Outcomes in Developing Countries: Evidence, Knowledge Gaps, and Policy Implications. Machin S, Woessmann L, Hanushek EA, editors. Handbook of the Economics of Education, 2016. 2016;653–743.

19. Baez Ramirez JE, Camacho A. What’s the long-term impact of conditional cash transfers on education? 2013 [cited 2025 Mar 28]; Available from: https://policycommons.net/artifacts/1505415/whats-the-long-term-impact-of-conditional-cash-transfers-on-education/2168491/

20. Austrian K, Maluccio JA, Soler-Hampejsek E, Muluve E, Aden A, Wado YD, et al. Long-term impacts of a cash plus program on marriage, fertility, and education after six years in pastoralist Kenya: A cluster randomized trial. SSM - Population Health. 2024 Jun 1;26:101663.

21. Girls Not Brides, UNFPA, Population Council and UNICEF. Cash and asset incentive schemes to address child marriage and support married girls [Internet]. 2024 Dec. Available from: https://www.girlsnotbrides.org/documents/2261/CRANK_Research_Spotlight_Cash_asset_incentive_schemes.pdf

